# State-Guided ICA of Functional Network Connectivity Reveals Temporal Signatures of Alzheimer’s Disease

**DOI:** 10.1101/2025.09.23.25336175

**Authors:** Elaheh Zendehrouh, Mohammad S. E. Sendi, Anees Abrol, Armin Iraji, Vince D. Calhoun

## Abstract

Identifying robust neuroimaging biomarkers for Alzheimer’s disease (AD) and mild cognitive impairment (MCI) is essential for early diagnosis and intervention. In this study, we introduce a novel, fully automated, guided dynamic functional connectivity (dFNC) framework for extracting multiple dynamic measures to distinguish MCI/AD from cognitively normal (CN) individuals.

Resting-state fMRI data were used to extract subject-specific brain networks via spatially constrained independent component analysis (scICA), using a multi-objective optimization framework to ensure alignment with known functional networks while preserving individual variability. Using these components, dFNC was computed through a sliding-window approach. ICA was then applied to the concatenated dFNC matrices from the UK Biobank (UKBB) dataset to identify five canonical brain states, each representing a replicable, independent pattern of connectivity. These states served as biologically informed priors in a state-constrained ICA (St-cICA), which was applied to each subject in the combined OASIS-3 and ADNI datasets to guide individual-level decomposition and ensure interpretable connectivity states guided by state priors derived from the normative UKBB sample.

St-cICA extracted subject-specific dFNC features and associated weighted timecourses. To characterize dFNC patterns, we computed metrics from the most strongly expressed (primary) state and introduced estimation of the second-most expressed (secondary) state at each time point, including dwell time, occupancy rate, and transition probabilities. Group comparisons using two-sample t-tests revealed widespread and significant alterations in AD/MCI compared to CN individuals. AD/MCI participants exhibited higher dwell times and increased self-transitions, indicating reduced neural flexibility and a tendency to remain in specific connectivity states. In contrast, CN individuals showed more diverse and recurrent transitions, reflecting greater adaptability. Secondary transitions revealed widespread selective switching in CN, whereas AD/MCI showed reduced cross-state engagement.

A classification model trained on 6,960 dynamic features achieved strong performance in distinguishing AD/MCI from CN (mean AUC ≈ 0.85). These findings highlight the potential of guided dFNC as a biomarker framework for early-stage AD detection using resting-state fMRI.

## 1. Introduction

Alzheimer’s disease (AD) is the most common cause of dementia worldwide, progressively eroding memory, executive function, and daily independence(1). Pathologically, AD disrupts large-scale brain networks long before the full clinical syndrome emerges, making neural connectivity an attractive target for early diagnosis and therapeutic monitoring (2–4). Mild cognitive impairment (MCI) captures this prodromal window, when timely intervention could meaningfully delay conversion to dementia(5, 6). Yet, current clinical assessments rely heavily on invasive or costly biomarkers —cerebrospinal fluid assays, amyloid PET —which may not be widely accessible (7, 8).

Notably, the widely accepted A/T/N classification system (9–11) emphasizes amyloid, tau, and neurodegeneration. Still, it omits neural dynamics, despite growing evidence that dynamic alterations in functional connectivity may precede structural decline or overt symptoms(12, 13). This presents a critical gap, as dynamic functional markers could offer earlier and more sensitive indicators of disease. Functional MRI (fMRI), a non-invasive and widely available technique that captures time-varying neural processes, provides a promising tool for addressing this gap (14, 15). The integration of dynamic fMRI metrics with established markers of neurodegeneration may enhance the early detection and monitoring of AD, representing an important direction for future research. (16–19).

Traditional fMRI analyses quantify static functional network connectivity (sFNC), a single correlation matrix that summarizes an entire resting-state scan (20). While intuitive, the sFNC model implicitly assumes that inter-regional communication is stationary, ignoring the brain’s rich temporal dynamics. Dynamic FNC (dFNC) lifts this assumption by capturing moment-to-moment fluctuations that encode how networks integrate and disengage in response to internal processing or spontaneous arousal changes (21–25). Early dFNC work has already linked AD to dwell times in low-integration states, characterized by longer dwell times and fewer transitions between states, as well as blunted temporal variability (12, 18, 26, 27). However, most studies derive these findings from group-level clustering: time windows from all participants are pooled, clustered, and then each subject’s temporal profile is back-projected(28). This “one-size-fits-all” strategy can obscure individual variability–a critical limitation given the clinical heterogeneity of AD(29)–and is especially vulnerable to noise stemming from short scan durations and low signal-to-noise ratios(30, 31). Moreover, aligning connectivity states across individuals is notoriously difficult; mismatched states can mask disease-relevant nuances.

To address these limitations, we developed a fully automated, state-constrained dFNC framework that unites two levels of spatially constrained independent component analysis (scICA) (32)with a novel state prior strategy. In Stage 1, we apply NeuroMark scICA (33) to resting-state fMRI, leveraging multi-objective optimization with reference (MOO-ICAR) to balance three goals: (i) maximize statistical independence of the extracted components; (ii) maintain high spatial similarity to template intrinsic connectivity networks (ICNs) derived from large normative samples; and (iii) preserve subject-specific variance essential for clinical phenotyping. This combination yields robust, biologically interpretable ICNs while attenuating noise and between-subject idiosyncrasies that plague unconstrained ICA.

Stage 2 targets temporal dynamics. We compute sliding-window dFNC for 1,000 healthy adults from the UK Biobank, apply ICA to these matrices, and distill five canonical connectivity states that dominate normative neural dynamics. These canonical states act as priors in a state-constrained ICA (St-cICA) applied to individual dFNC data from OASIS-3 and ADNI. By “nudging” decomposition toward well-characterized state templates, we enforce cross-subject comparability without erasing clinically meaningful deviations (34, 35). Next, we implement state-wise mean removal: each time point is assigned to its nearest canonical state via Euclidean distance, the subject-specific mean pattern within that state is subtracted, and the residual captures transient excursions around a personalized baseline. This correction boosts sensitivity to subtle fluctuations–critical under the low-signal to noise (SNR) conditions common in clinical fMRI– while suppressing static background structure. Because St-cICA returns continuous state-engagement time courses, our framework naturally accommodates partial occupancy and smooth transitions, thereby avoiding the artificial discreteness of hard clustering (34). Importantly, every step is fully automated, making the pipeline scalable to biobank-scale data and replicable across sites.

Grounded in prior evidence of network rigidity in AD, we hypothesize that patients with MCI or AD will (i) linger longer in low-integration states, (ii) allocate a larger fraction of scanning time to such states, and (iii) transition between states less frequently and less flexibly than cognitively normal (CN) peers. Spatially, we expect heightened expression of hypo-connected patterns and diminished engagement of highly integrated states, reflecting breakdowns in distributed communication. These signatures should manifest as prolonged dwell times, skewed occupancy rates, and suppressed transition probabilities–metrics that index the brain’s capacity for flexible reconfiguration (13, 26, 27, 36) By capturing these individualized dynamics, our framework aims to deliver a practical dFNC-based biomarker set that can stage cognitive impairment, stratify patients for clinical trials, and ultimately track treatment response across the Alzheimer’s disease continuum.

## 2. Materials and Methods

### 2.1. Datasets and study population

This study draws on three large-scale neuroimaging datasets selected to support different stages of analysis: the UK Biobank or UKBB (37), the Open Access Series of Imaging Studies or OASIS-3 (38), and the Alzheimer’s Disease Neuroimaging Initiative or ADNI (39). Together, these datasets span cognitively normal (CN), mild cognitive impairment (MCI), and Alzheimer’s disease (AD), offering complementary strengths for constructing and validating dynamic connectivity models.

We leveraged resting-state fMRI data (5 minutes) from 1,000 healthy individuals in the UK Biobank to derive canonical dFNC states. Independent component analysis (ICA) was applied to their dFNC matrices to extract these states, which were subsequently used as spatial priors in downstream constrained ICA analyses(37). The OASIS-3 resting-state fMRI data (6 minutes) contain 1,389 imaging samples (mean age: 67.18 ± 8.71) along with demographic and clinical data (38). To distinguish MCI from cognitively normal individuals, we applied the Clinical Dementia Rating Sum of Boxes (CDR_SOB) scale, with MCI defined as CDR_SOB > 0 and CN as CDR_SOB = 0(40). This resulted in 1,028 CN samples and 361 MCI samples. Additionally, we included the ADNI resting-state fMRI data (5 minutes) (mean age: 71.85 ± 6.99), which comprises 488 CN, 229 MCI, and 99 AD individuals. ADNI participants were classified based on CDR scores, with CN defined as CDR = 0, MCI as CDR = 0.5, and AD as CDR = 1(41). **Table 1** presents the demographic information for the OASIS-3 and ADNI datasets.

**Table 1.**
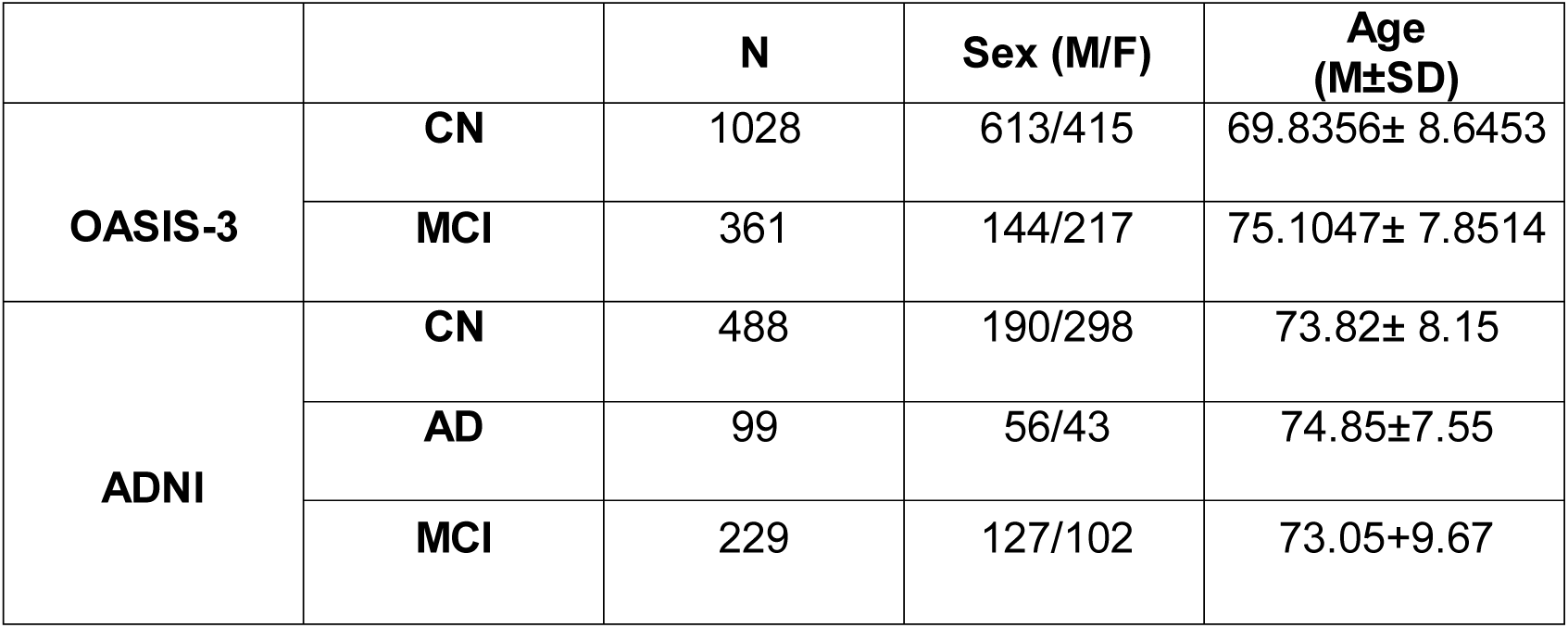
Demographic and clinical information of OASIS-3 and ADNI datasets.

### 2.2. Imaging protocol

UKBB data were collected on 3T Siemens scanners equipped with 32-channel head coils, using a gradient-echo echo planar imaging (EPI) sequence with a resolution of 2.4 × 2.4 × 2.4 mm³, echo time (TE) of 39 ms, repetition time (TR) of 735 ms, and a field-of-view of 88 × 88 × 64. OASIS-3 data were acquired using three Siemens scanners: a 1.5T Vision scanner with a 16-channel head coil and two 3T TIM Trio scanners with 20-channel head coils, using a gradient-echo EPI sequence. Imaging parameters for the 3T Trio scanners included TE = 27 ms, TR = 2.5 s, flip angle = 90°, slice thickness = 4 mm, and matrix size = 64 × 64, while the 1.5T Vision scanner had a TR of 2.2 s. The ADNI dataset included imaging from multiple scanner manufacturers, including 1.5T and 3T GE, Philips, and Siemens systems. During data collection, participants had their eyes open. The minimum TR in the ADNI dataset is 0.6 seconds, and we used a sliding window of 60 TRs for dynamic connectivity analysis, corresponding to a window size of 36 seconds (39).

### 2.3. Preprocessing and feature extraction

Preprocessing of resting-state fMRI data was performed using Statistical Parametric Mapping (SPM12). The first five dummy scans were removed before slice timing correction, with the middle slice used as a reference. Rigid body motion correction was applied to correct for participant head movement, and the images were normalized to Montreal Neurological Institute (MNI) space using an echo-planar imaging template. A Gaussian kernel with a full width at half maximum (FWHM) of 6 mm was applied for spatial smoothing.

Independent components (ICs) were extracted using version 1.0 of the NeuroMark framework, a spatially constrained ICA method that leverages ICA component maps derived from two large-scale (N>900) datasets, the Human Connectome Project (HCP) and the Genomics Superstruct Project (GSP) (33). NeuroMark has been validated in multiple studies and provides a standardized template for identifying functionally relevant networks. This framework decomposes fMRI data into 53 ICs, organized into seven functional networks: subcortical network (SC), auditory network (AUD), sensorimotor network (SM), visual network (VS), cognitive control network (CC), default mode network (DM), and cerebellar network (CB) as shown in **Figure 2**. The NeuroMark templates were applied using the Group ICA for fMRI Toolbox GIFT (42) , with additional preprocessing including detrending (linear, quadratic, cubic), multiple regression to remove motion confounds, despiking to eliminate outliers, and low-pass filtering at 0.15 Hz. This pipeline has been used in numerous previous dFNC studies (10, 43–45).

**Figure 1.**
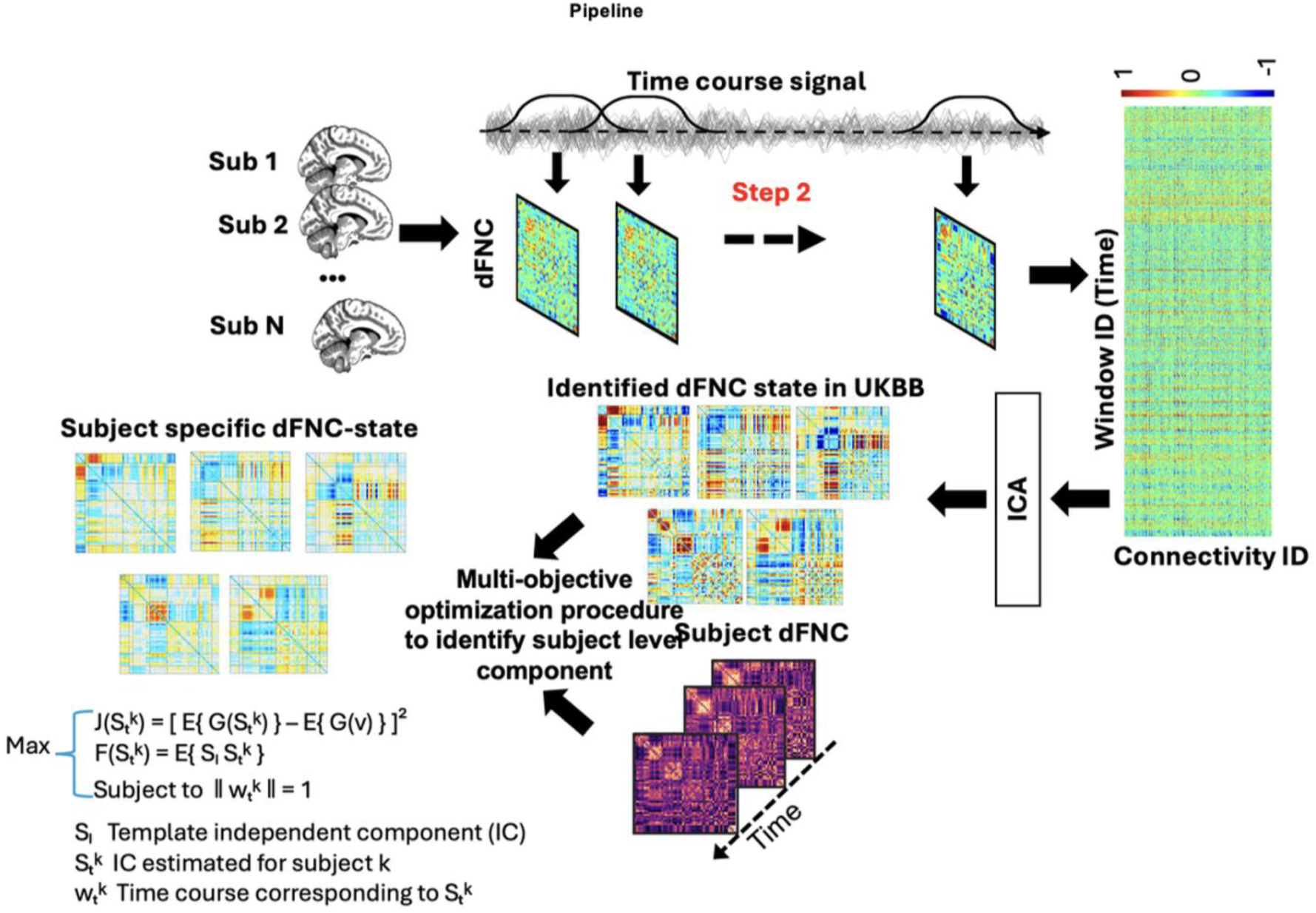
Workflow for identifying dFNC patterns in MCI/AD and CN groups. In Step 1, resting-state fMRI data were collected for individual subjects across multiple datasets. Step 2: Time course signals were extracted to compute dFNC matrices, capturing time-varying connectivity. Step 3: ICA was performed on UKBB data to identify five canonical brain states (State 1 to State 5). Step 4: Constrained FNC analysis was applied to the combined datasets using these reference states, enabling consistent subject-level dFNC comparisons across groups.

**Figure 2.**
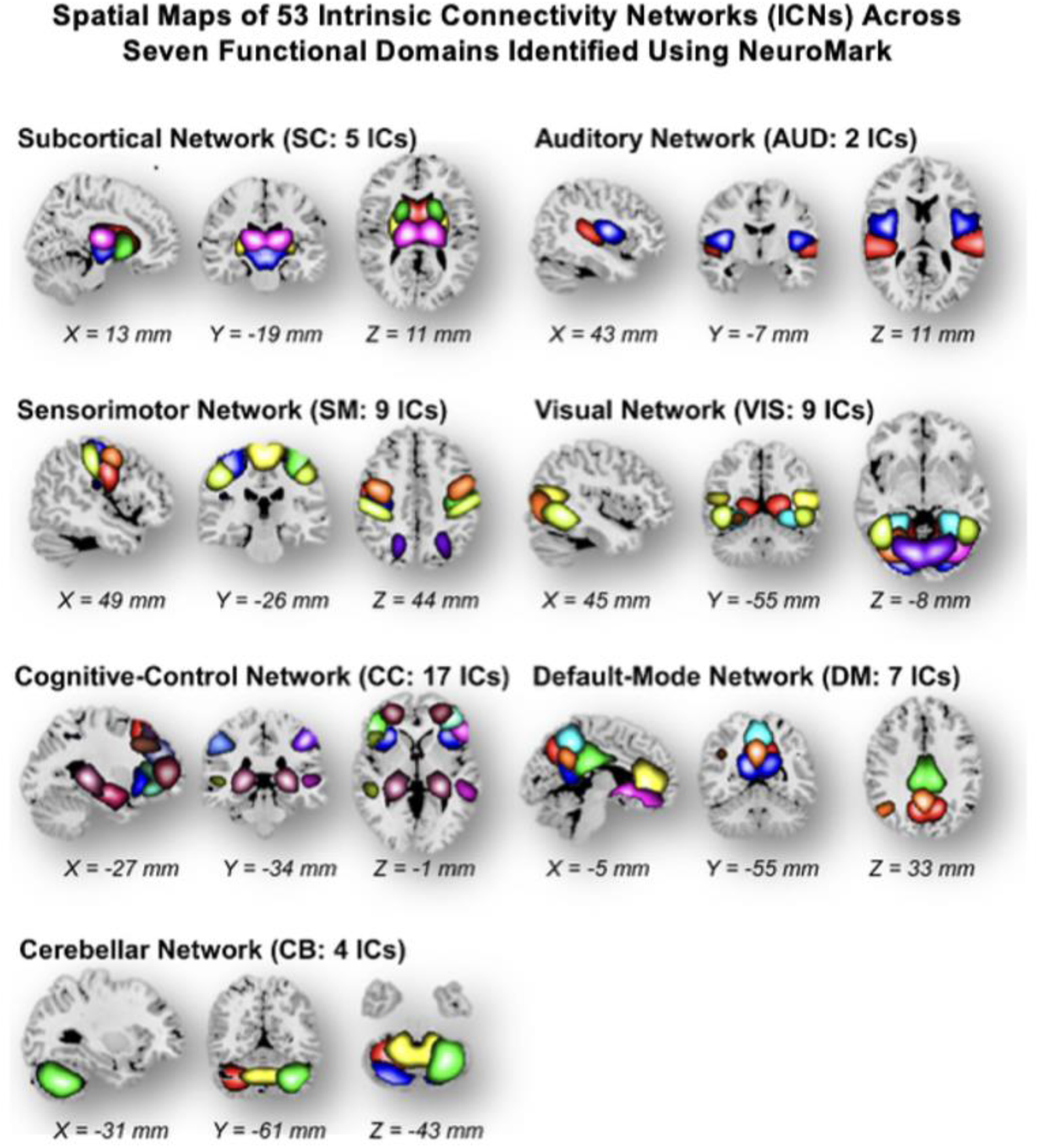
NeuroMark pipeline was employed to extract 53 robust intrinsic connectivity networks (ICNs) from resting-state fMRI data, categorized into seven functional domains: subcortical (SC, 5 ICs), auditory (AUD, 2 ICs), sensorimotor (SM, 9 ICs), visual (VIS, 9 ICs), cognitive control (CC, 17 ICs), default mode (DM, 7 ICs), and cerebellar (CB, 4 ICs). ICNs were identified by performing group ICA with a model order of 100 on two large healthy control datasets—Human Connectome Project (HCP) and Genomics Superstruct Project (GSP). Reproducible components were selected by spatial correlation (r > 0.4) and confirmed based on timecourse similarity. The resulting network templates provided biologically meaningful priors for downstream spatially constrained ICA and dynamic functional connectivity analyses.

### 2.4. Dynamic functional network connectivity

Dynamic FNC was estimated using a sliding window approach using the GIFT toolbox (46). A tapered window was constructed by convolving a rectangular window with a Gaussian kernel (σ = 3). Within each window, Pearson correlations were computed among 53 ICs estimated from NeuroMark, yielding a three-dimensional dFNC array (C × C × T), where C = 53 and T represents the number of windows (450 for UKBB, 139 for OASIS-3, and 448 for ADNI).

### 2.5. Implementing ICA and constrained ICA

To identify brain states in the combined dataset, we first estimated canonical brain states in the UKBB dataset using FNC-ICA. This decomposes time-varying connectivity into independent connectivity patterns, allowing for the identification of recurrent states. The input matrix consisted of cell-by-window representations of network connectivity, where each “cell” corresponds to a specific functional network connection, and each “window” captures a short temporal segment over which functional connectivity is estimated. We then performed ICA on the concatenated windowed dFNC matrices from the UKBB dataset, using principal component analysis (PCA) to reduce dimensionality to five canonical states. ICA was applied to the whitened data using the Infomax algorithm via the ICATB toolbox, yielding spatially independent components that reflect canonical brain states.

These states represented the most recurrent and representative connectivity configurations observed in the UKBB healthy control population. Using these predefined states from UKBB, constrained ICA was applied to the combined OASIS-3 and ADNI dataset to extract subject-specific connectivity states. Each subject’s data was decomposed into window-by-state and state-by-FNC matrices. To ensure comparability across subjects, state calibration was performed using an ICA-based regression approach, where subject-specific dFNC data were regressed onto the group-level ICA components to estimate beta weights (47). These weights were then used to reconstruct and scale individual state expressions, normalizing both spatial and temporal features across individuals.

To further refine our dFNC analysis, we applied a state-wise mean removal approach. Each timepoint was assigned to a state based on Euclidean distance to the precomputed state means. Within each state, the mean dFNC was calculated and subtracted from all assigned timepoints, preserving dynamic fluctuations while removing static components. This approach allowed for more accurate characterization of transient network dynamics. The corrected dFNC data were then analyzed using constrained ICA to extract stable connectivity patterns.

### 2.6. Primary and secondary dwell-times, occupancy rate, and state transition probability

After identifying subject-specific dFNC in our target datasets, including OASIS-3 and ADNI, we extracted latent features for further analysis. Prior studies have quantified individual engagement with dynamic brain states using metrics such as dwell time, occupancy rate, and state transition probability (48). To compute these features, we first generated state vectors for each participant by comparing each time-resolved dFNC matrix to a set of canonical dFNC states using Euclidean distance.

In our framework, we constructed two distinct state vectors per participant. The primary state vector assigns to each time point the closest canonical dFNC state–i.e., the one with the smallest Euclidean distance–reflecting dominant state dynamics. In addition, we introduce a novel secondary state vector, which assigns the second-closest state to each time point based on the following smallest distance. While the primary state vector captures dominant patterns of functional connectivity, this secondary vector enables the investigation of subdominant and transitional state dynamics that may otherwise be obscured.

By modeling second-state properties, we obtain a richer and more sensitive view of brain flexibility, partial state engagement, and transient dynamic transitions. These second-level dynamics may be particularly relevant for detecting subtle disruptions in neurodegenerative conditions such as Alzheimer’s disease (AD), where early impairments in the brain’s ability to reconfigure flexibly may precede large-scale network reorganization. They also offer additional mechanistic insights into the brain’s capacity to reallocate functional resources in response to internal or external demands.

From both state vectors, we derived three core dynamic features: dwell time, occupancy rate, and state transition probability. Dwell time represents the average number of consecutive time points spent in a given state before switching. Occupancy rate reflects the overall proportion of time spent in each state. State transition probability quantifies the likelihood of transitioning from one state to another over time. Specifically, first and second dwell times were calculated as the mean durations spent in the most frequently assigned state and the second-most frequently assigned state, respectively(49).

These dynamic metrics were computed separately from the primary and secondary state vectors, yielding primary and secondary measures of dwell time, occupancy rate, and transition probability. Group-level differences between cognitively normal (CN) and AD/MCI participants were assessed using two-sample *t*-tests to determine whether these features could differentiate healthy from impaired dynamic brain function.

### 2.7. Classification

To classify AD/MCI from CN, we employed elastic net regularization (ENR)-based feature selection combined with 5-fold cross-validation (50). A total of 6,960 features were used for classification, including 6,890 brain connectivity features (1,378 features across each of the five states), 10 features representing first and second dwell times, 10 features for first and second occupancy rates, and 50 features capturing first and second state transition probabilities. To evaluate classifier robustness, we implemented a randomized resampling procedure over ten iterations, where in each iteration, 689 AD/MCI subjects were randomly paired with 689 healthy CNs. The model was trained and evaluated on each newly formed dataset, and area under the curve (AUC) values were recorded to assess performance stability(51). The ENR feature learning method was used to identify the top 50 predictors in the classification between AD/MCI from CN. This methodology allowed for the identification of dFNC alterations associated with AD and MCI, providing new insights into brain state transitions and their role in neurodegeneration. By leveraging a constrained ICA-based approach with state-wise mean removal, this study enhances the ability to capture subtle changes in FNC dynamics that may serve as early biomarkers for AD(52).

## 3. Results

### 3.1. Identification of canonical brain states in the UKBB dataset

To establish a reference set of FNC brain states, we applied FNC-ICA to the UKBB dataset. This analysis identified five distinct connectivity states, representing recurring patterns of FNC in a large sample of cognitively normal individuals. These canonical brain states serve as a normative reference and will be used to assign subject-specific dFNC states in our target datasets, including OASIS-3 and ADNI. Figure 3 presents the extracted five canonical brain states from UKBB, where each matrix illustrates the connectivity pattern of a given state. These states display distinct patterns of inter-network connectivity across seven ICNs (33, 48, 53). Each state reveals unique connectivity dynamics among these networks, with variations in the strength and organization of functional connections.

**Figure 3.**
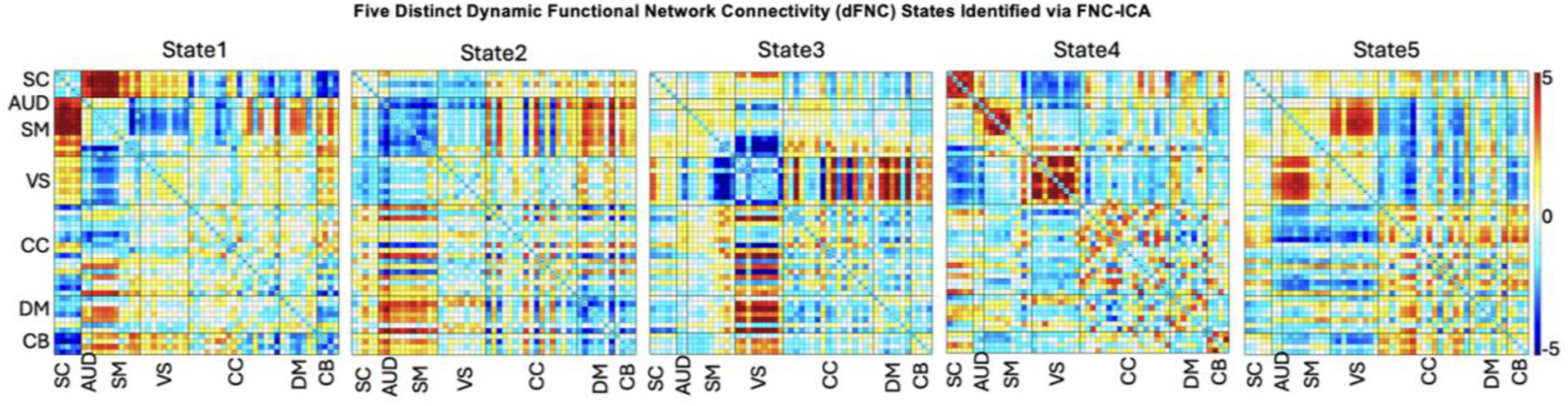
Maximally Independent Brain States Identified through FNC ICA. Five distinct brain states are presented, representing the average connectivity patterns in each state across subjects. The connectivity matrices display correlations between brain regions categorized as SC, AUD, VS, CC, DM, and CB. The color scale ranges from -5 to 5, indicating the strength and direction of connectivity. State 1 to State 5 highlight unique functional network connectivity patterns, providing insight into dynamic brain state changes and their implications for cognitive function.

State 1 exhibits strong positive connectivity among SC with SM and AUD network regions, while interactions between CC with AUD and SM networks are notably reduced. State 2 presents a more uniform, globally distributed pattern of weak inter-network connectivity, often referred to as a “flattened” state in previous literature(48), compared to the strongly modular or segregated profiles seen in other states. Also, presents a flattened connectivity pattern with weakly connected network interactions, suggesting a more distributed functional state (21). State 3 is characterized by greater negative connectivity within and between sensory networks. Additionally, it exhibits increased connectivity between the VIS and other brain networks, including the CC, DM, and CB networks. States 4 and 5, however, reveal a contrasting scenario with increased connectivity within and between the SM and VS networks, suggesting enhanced interaction and coordination between these regions. State 4 further exhibits more pronounced positive connectivity within the SC and CB networks, indicating a robust link between these regions that may support complex motor functions.

Additionally, State 5 not only shows enhanced connectivity within the CC network but also exhibits substantial positive connections between the CC, DM, and CB networks. This extended connectivity suggests a dynamic integration of cognitive control, introspective or resting state functions, and cerebellar processing. These diverse connectivity patterns across different brain states provide insights into the variable functional brain pattern and their potential implications for cognitive and motor functions in different physiological or pathological states. This extended connectivity suggests a dynamic integration of cognitive control, introspective or resting state functions, and cerebellar processing (22, 30). These diverse connectivity patterns across different brain states provide insights into the variable functional brain pattern and their potential implications for cognitive and motor functions in various physiological or pathological states. In the next step, these states serve as the functional priors for the constrained ICA applied to the OASIS-3 and ADNI datasets, allowing for a biologically informed decomposition of subject-specific brain connectivity dynamics (54).

### 3.2. Group differences in dynamic functional connectivity patterns

After applying constrained ICA to estimate subject-specific dFNC states in the OASIS-3 and ADNI datasets, we examined group differences in functional connectivity across the five identified states. Two-sample *t*-tests were performed for each connection, and false discovery rate (FDR) correction was applied to account for multiple comparisons. The analysis revealed significant connectivity differences between CN and MCI/AD individuals, as illustrated in Figure 4. In this figure, the lower triangle displays connections that were significant before FDR correction, while the upper triangle highlights those that remained significant after correction. Across the five states, distinct dFNC differences emerged between the CN and patient groups. In State 1, the CN group exhibited stronger connectivity between the AUD, SM, and VIS networks with the SC network, whereas the MCI/AD group showed stronger connectivity between the SC network and the CC, DM, and CB networks. Additionally, in the CN group, the AUD and SM networks demonstrated stronger connectivity with the CB network. In State 2, there was significantly stronger SC–CB connectivity in the CN group, while the DM–CB connectivity was significantly higher in the MCI/AD group. These alterations in SC–CB interactions may reflect compensatory mechanisms or disruptions in functional integration in CN. State 3 revealed significant differences in connectivity between the VIS and CC networks, with both positive and negative shifts observed in CN compared to MCI/AD individuals. The DM and CB networks also showed higher connectivity in CN, suggesting disruptions in higher-order cognitive processing associated with neurodegeneration. In State 4, CN individuals exhibit stronger connectivity between the SC and CB networks, whereas MCI/AD individuals show stronger connectivity between the VIS and CB networks. State 5 reveals significant differences in connectivity within and between the AUD, SM, and VIS networks, with CN individuals showing stronger connectivity than MCI/AD individuals in these sensory domains. Conversely, MCI/AD individuals exhibit stronger connectivity between the SM and CB networks. These alterations in sensorimotor-cerebellar interactions may reflect compensatory mechanisms or disruptions in motor coordination associated with neurodegeneration.

**Figure 4.**
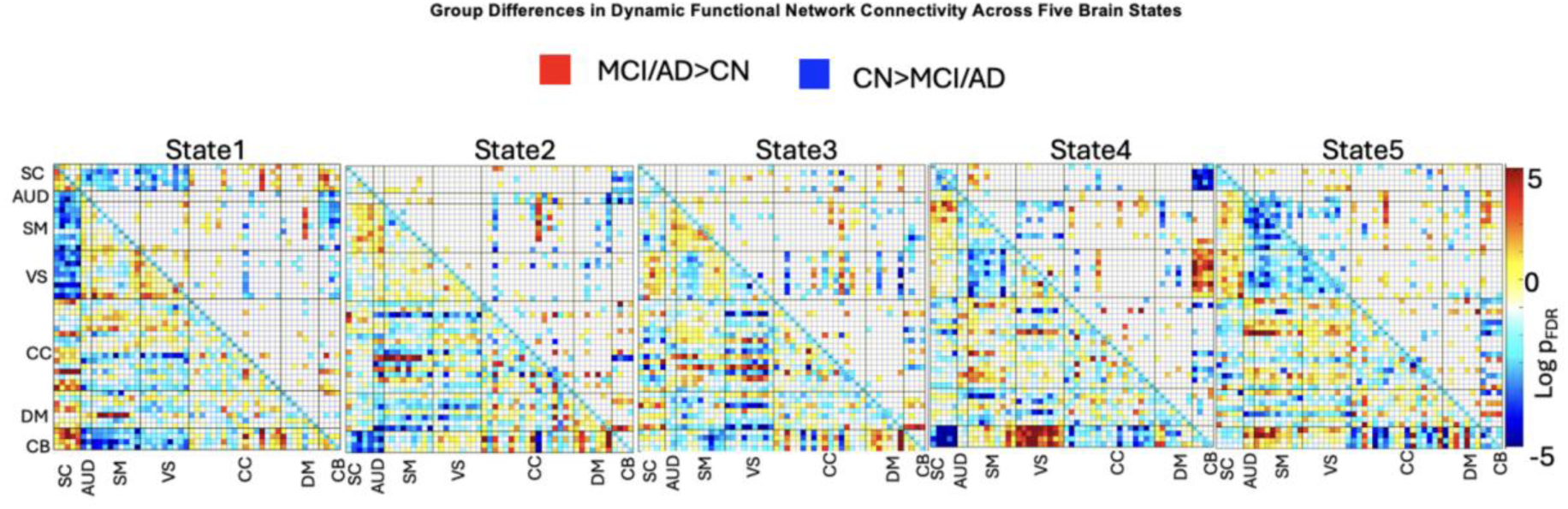
Two-sample t-tests comparing CN individuals to a merged group of MCI and AD participants across five dynamic functional connectivity states. The upper triangle of each matrix shows connections with significant group differences after FDR correction. Red indicates stronger connectivity in MCI/AD, while blue indicates stronger connectivity in CN. Each subplot corresponds to a distinct brain state, illustrating spatially specific functional connectivity alterations linked to cognitive impairment and neurodegeneration.

### 3.3. Group differences in dwell time across states

To further investigate dFNC patterns, we evaluated dwell time metrics based on the strongest (i.e., primary dwell time) and second strongest (i.e., secondary dwell time) connectivity states. At each time point, a subject’s primary dwell time corresponds to the most strongly expressed state, while the secondary dwell time represents the next most engaged state. This distinction allows us to assess both primary state stability and secondary state transitions in CN and MCI/AD individuals.

**Figure 5. A** illustrates the comparison of mean primary dwell times between the CN group and the MCI/AD. The MCI/AD group exhibited significantly longer dwell times across all five states (p < 0.05, indicated by asterisks), suggesting a reduced ability to transition between dFNC states. In contrast, the CN group had shorter dwell times, reflecting greater neural flexibility and a higher frequency of transitions between connectivity patterns. The two-sample t-test results revealed significant group differences in mean primary dwell times of all states. In State 1, the mean primary dwell time was 15.56 for the CN group and 17.14 for the MCI/AD group (FDR p = 0.01). In State 2, the CN group had a primary dwell time of 13.52, while the MCI/AD group exhibited 15.97 (FDR p = 3.99 × 10^-5^). In State 3, the CN group exhibited a primary dwell time of 15.63, compared to 18.82 in the MCI/AD group (FDR p = 1.05 × 10^-6^). Similarly, in State 4, the CN group’s primary dwell time was 16.61, while the MCI/AD group remained in the state for 18.25 (FDR p = 0.01). In State 5, the CN group exhibited a primary dwell time of 14.71, whereas the MCI/AD group had 16.99 (FDR p = 1.91 × 10^-4^). These findings indicate that MCI/AD individuals tend to remain in specific FNC states for extended durations, suggesting increased network rigidity and reduced neural flexibility. The prolonged primary dwell times in the MCI/AD group may reflect difficulties in dynamically reconfiguring brain networks, which could contribute to cognitive impairments observed in neurodegenerative conditions.

**Figure 5.**
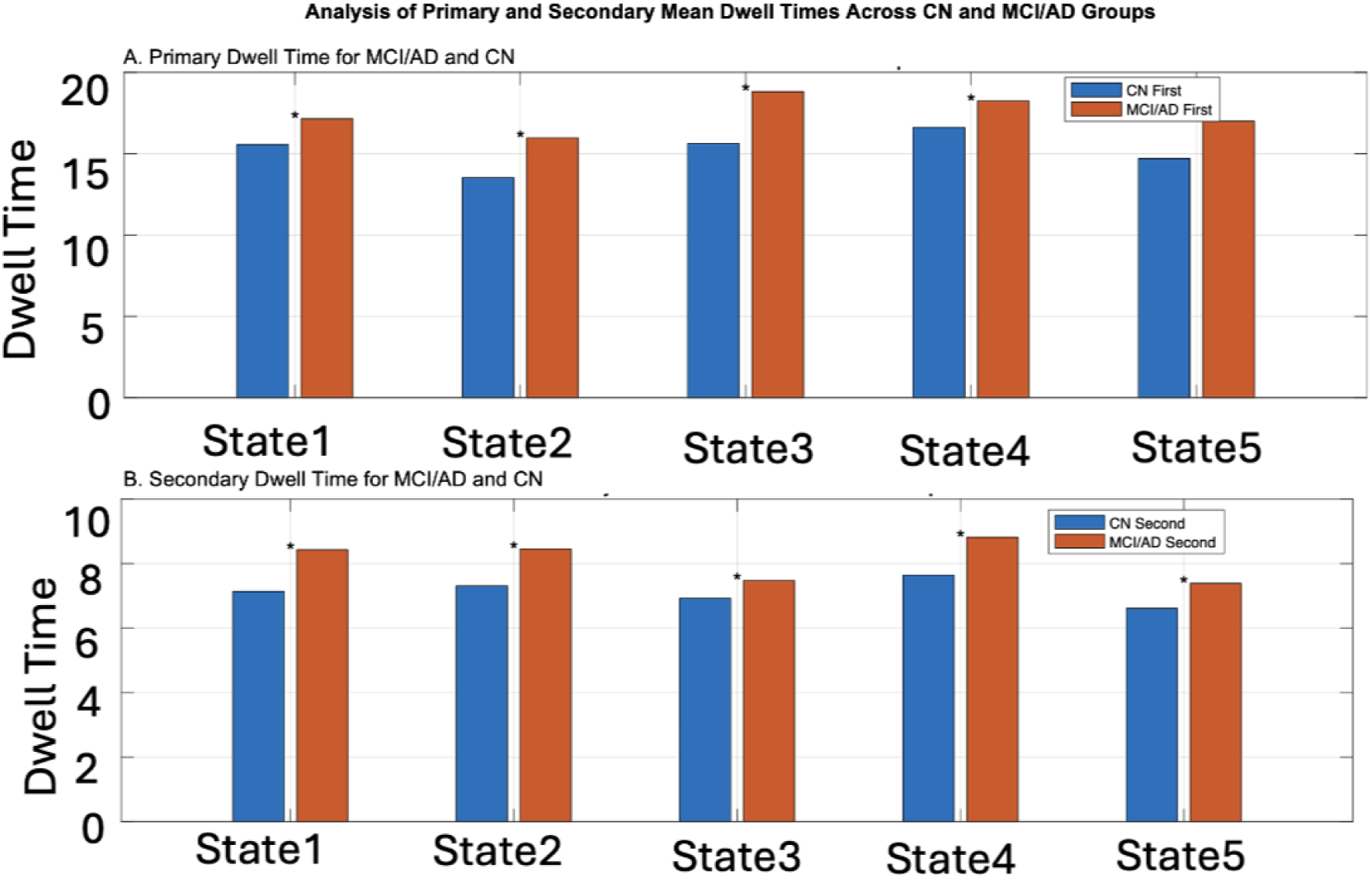
(A–B) Primary and Secondary Dwell Times for CN and AD/MCI Groups. Bar graphs show primary (top) and secondary (bottom) dwell times for CN and AD/MCI groups across brain states. Significant differences (FDR-adjusted p < 0.05) are marked with asterisks (*), indicating longer dwell times in the AD/MCI group across all states in both primary and secondary methods.

**Figure 5. B** presents the mean secondary-dwell time comparisons between the CN and MCI/AD groups. Similar to the first dwell time analysis, the MCI/AD group exhibited significantly longer secondary dwell times across all five states (p < 0.05), suggesting reduced flexibility not only in their dominant states but also in their secondary network states.

The two-sample t-test results showed that in State 1, the mean secondary dwell time was 7.14 for the CN group and 8.43 for the MCI/AD group (FDR p = 4.99 × 10^-7^). In State 2, the CN group had a secondary dwell time of 7.31, while the MCI/AD group showed 8.45 (FDR p = 7.59× 10^-7^). In State 3, the CN group exhibited a secondary dwell time of 6.92, compared to 7.47 in the MCI/AD group (FDR p = 0.0057). Similarly, in State 4, the CN group’s secondary dwell time was 7.64, while the MCI/AD group remained in the state for 8.82 (FDR p = 4.99 × 10^-7^). In State 5, the CN group exhibited a secondary dwell time of 6.61, whereas the MCI/AD group had 7.38 (FDR p = 2.68 × 10^-4^). These results suggest that MCI/AD patients not only exhibit prolonged engagement in their dominant states but also reduced flexibility in their secondary states, potentially indicating compensatory mechanisms or impaired adaptive transitions (26, 55). In contrast, CN individuals demonstrate greater adaptability, as evidenced by more frequent transitions between primary and secondary connectivity states. The increased dwell time stability in MCI/AD individuals reinforces the hypothesis that reduced dynamic network flexibility may underlie cognitive decline in neurodegenerative diseases.

### 3.4. Group differences in occupancy rate

To further examine the dynamic characteristics of dFNC states, we analyzed occupancy rates in both primary and secondary states. Occupancy rate is defined as the proportion of time a subject spends in each state relative to the total scan duration. This measure provides insight into how frequently each group engages in specific FNC patterns. Figure 6 compares the mean primary occupancy rates between the CN and MCI/AD across the five states. The CN group exhibited a significantly higher primary occupancy rate in State 1 (CN: 0.22, MCI/AD: 0.21, FDR p = 4.91 × 10^-4^), suggesting that healthy individuals spend more time in this dFNC state. Conversely, the MCI/AD group demonstrated a significantly higher primary occupancy rate in State 3 (CN: 0.20, MCI/AD: 0.22, FDR p = 1.84 × 10^-4^), indicating a shift in network engagement in MCI/AD individuals. The primary occupancy rates for States 2, 4, and 5 did not show significant differences between groups, suggesting similar engagement in these connectivity patterns (State 2: CN = 0.17, MCI/AD= 0.18, FDR p = 0.33; State 4: CN = 0.1791, MCI/AD = 0.1733, FDR p = 0.13; State 5: CN = 0.2084, MCI/AD = 0.2108, FDR p = 0.45). These findings suggest that CN individuals tend to remain in State 1, dFNC state, while MCI/AD patients engage more frequently in State 3, potentially reflecting pathological network reorganization. The altered occupancy patterns in the patient group may indicate difficulties in maintaining typical dFNC states and an increased tendency to engage in alternative connectivity configurations associated with neurodegeneration (55).

**Figure 6.**
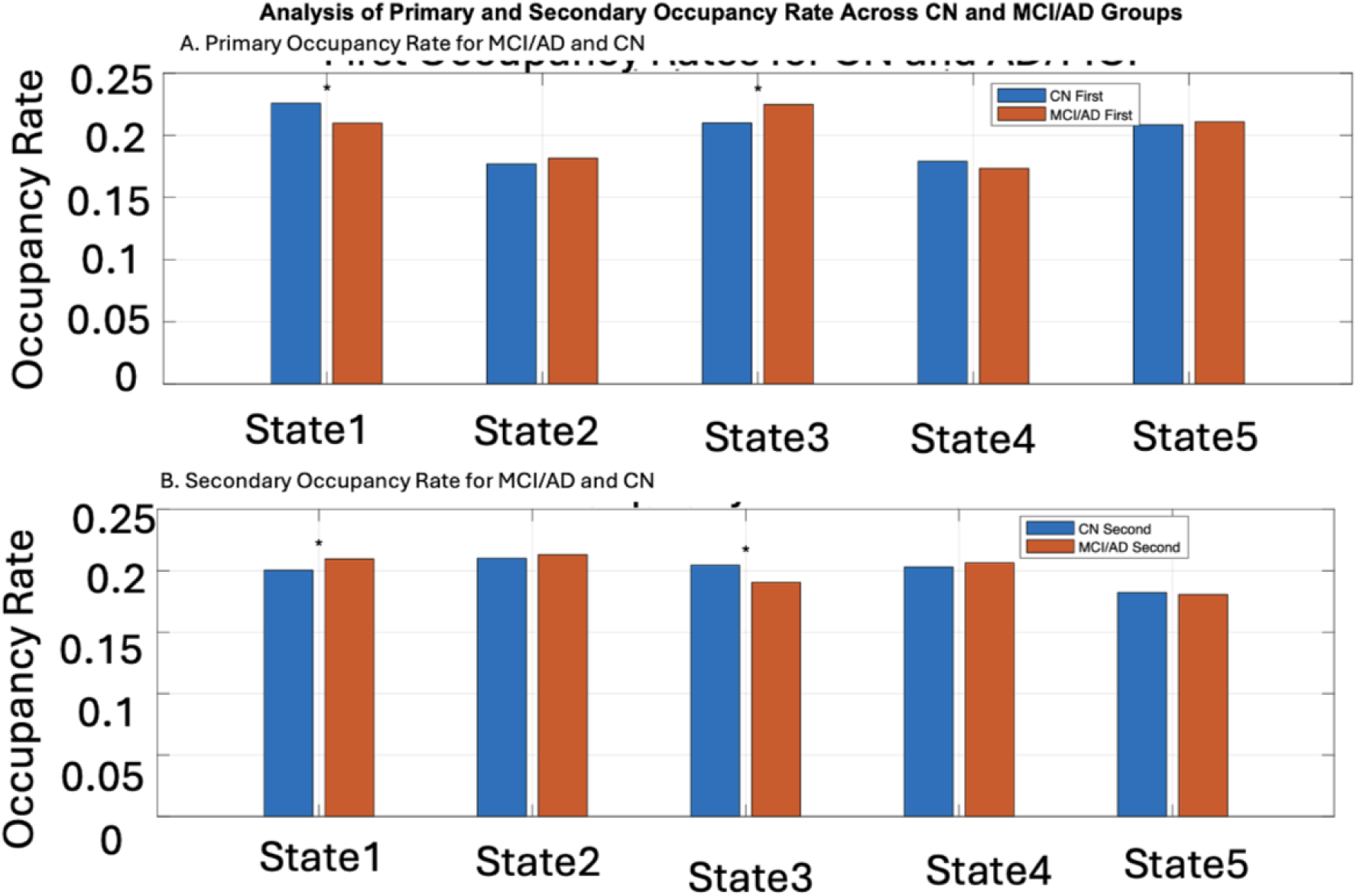
(A–B) Primary and Secondary State Occupancy Rates for CN and AD/MCI Groups. Bar graphs illustrate first (top) and second (bottom) occupancy rates for CN and AD/MCI groups across brain states. Significant differences (FDR-adjusted p < 0.05) are marked with asterisks (*).

**Figure 6.B** presents the comparison of mean secondary occupancy rates across states between the CN and MCI/AD groups. The MCI/AD group exhibited significantly lower occupancy rates in State 1 (CN: 0.22, MCI/AD: 0.21, FDR p = 0.03) and higher secondary engagement in State 3 (CN: 0.20, MCI/AD: 0.19, FDR p = 6.03×10^-4^) compared to the CN group. The remaining states did not show statistically significant differences, suggesting similar engagement patterns across the two groups. These results further indicate that MCI/AD patients exhibit altered engagement patterns, spending less time in certain dFNC states compared to healthy individuals. The increased occupancy in State 3 for CN individuals may reflect greater network flexibility, while the reduced engagement in this state by MCI/AD patients may suggest disruptions in network stability linked to neurodegeneration (26). The overall altered occupancy patterns in the MCI/AD group may indicate compensatory mechanisms or dysfunctional transitions, contributing to cognitive impairment in MCI/AD.

### 3.5. Group differences in state transition probabilities

To further characterize the temporal dynamics of FNC, we analyzed state transition probabilities, which quantify the likelihood of switching from one connectivity state to another. We examined both primary and secondary state transition probabilities to capture primary and secondary shifts in network dynamics, providing a more comprehensive assessment of neural flexibility and state stability.

**Figure 7. A** illustrates group-level differences in primary transition probabilities between individuals with CN and those with MCI/AD. Blue arrows denote transitions that were significantly more frequent in the CN group, while red arrows reflect transitions that were more frequent in MCI/AD. The thickness of each arrow represents the magnitude of the group difference, with statistical significance determined using FDR correction (p < 0.05 and p < 0.01). CN individuals exhibited more frequent transitions across distinct states–including between States 2, 3, 4, and 5–suggesting preserved flexibility and dynamic reconfiguration of network states. In contrast, MCI/AD individuals showed significantly elevated self-transitions (loops), indicating prolonged occupancy within specific states and reduced capacity to switch between functional configurations flexibly. These findings highlight reduced transition specificity and increased rigidity in MCI/AD, consistent with early-stage neurodegeneration.

**Figure 7.**
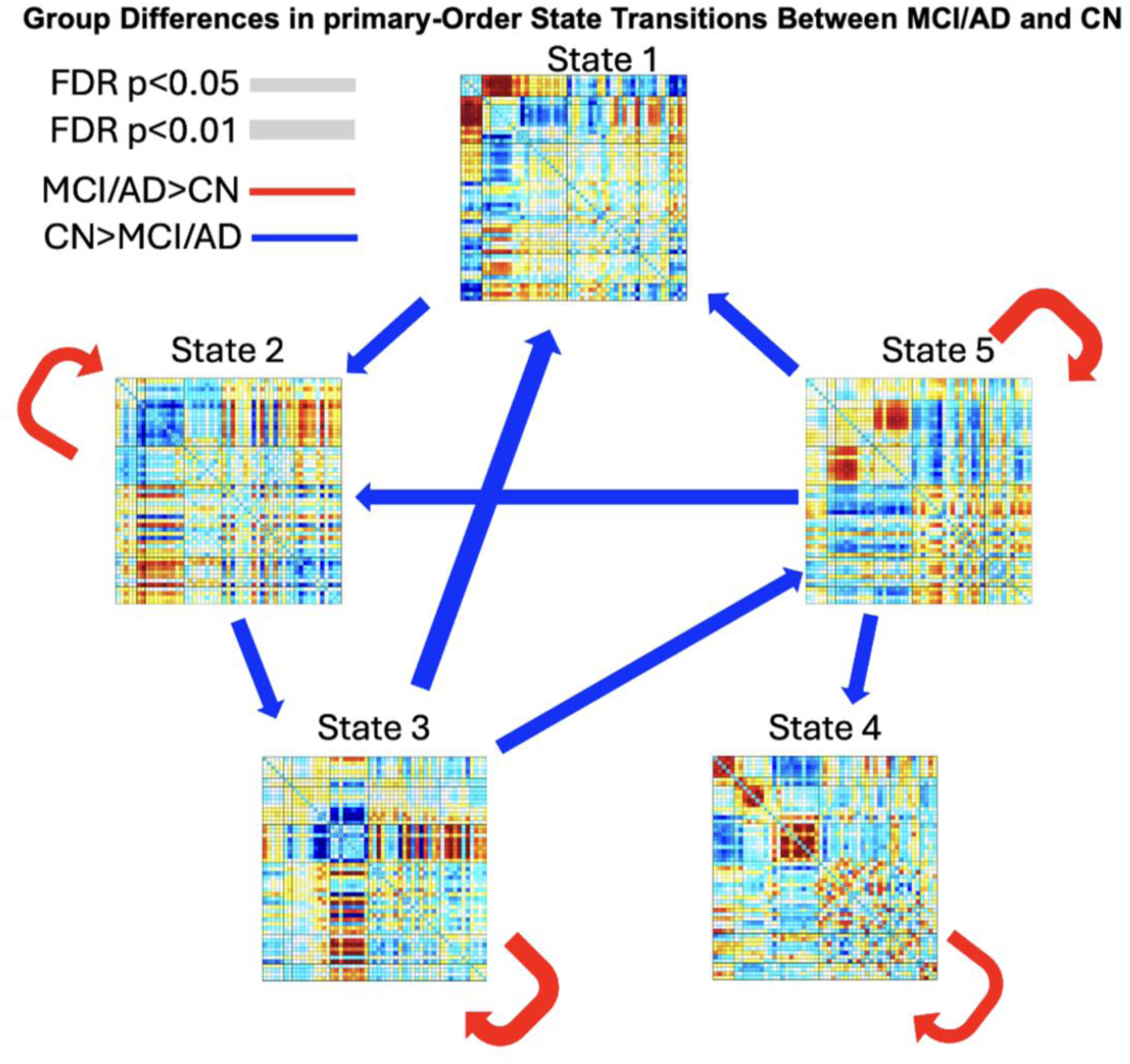
(A) Group-level differences in state transition probabilities between CN and MCI/AD individuals. Five canonical dFNC states are depicted, with directional arrows indicating the transitions between them. Red arrows: transitions significantly higher in MCI/AD; blue arrows: transitions higher in CN. Arrow thickness reflects effect size; significance is FDR-corrected (p < 0.05 and p < 0.01). CN individuals exhibited stronger recurrent transitions (loops) within States 2–5, suggesting preserved network flexibility, while MCI/AD showed elevated transitions toward more fragmented patterns (e.g., State 1), reflecting impaired network switching.

**Figure 7.B** Group-level differences in secondary state transition probabilities between cognitively normal (CN) and MCI/AD individuals. Arrows indicate statistically significant differences in the second most probable transitions. Blue arrows represent transitions that were more frequent in CN than in MCI/AD, while red arrows denote transitions that were more frequent in MCI/AD than in CN. Arrow thickness reflects the magnitude of group differences, with significance determined by FDR correction thresholds (*p* < 0.05 and *p* < 0.01). CN individuals demonstrated widespread increases in second-order transitions across nearly all state pairs, indicating greater temporal flexibility and dynamic coordination of brain networks. In contrast, MCI/AD individuals exhibited stronger self-transitions (loops), suggesting prolonged persistence within specific states and a reduced capacity for adaptive reconfiguration. These findings reflect diminished neural adaptability and a loss of transition richness in MCI/AD, consistent with impaired temporal dynamics and early neurodegenerative processes.(22).

**Figure 7.**
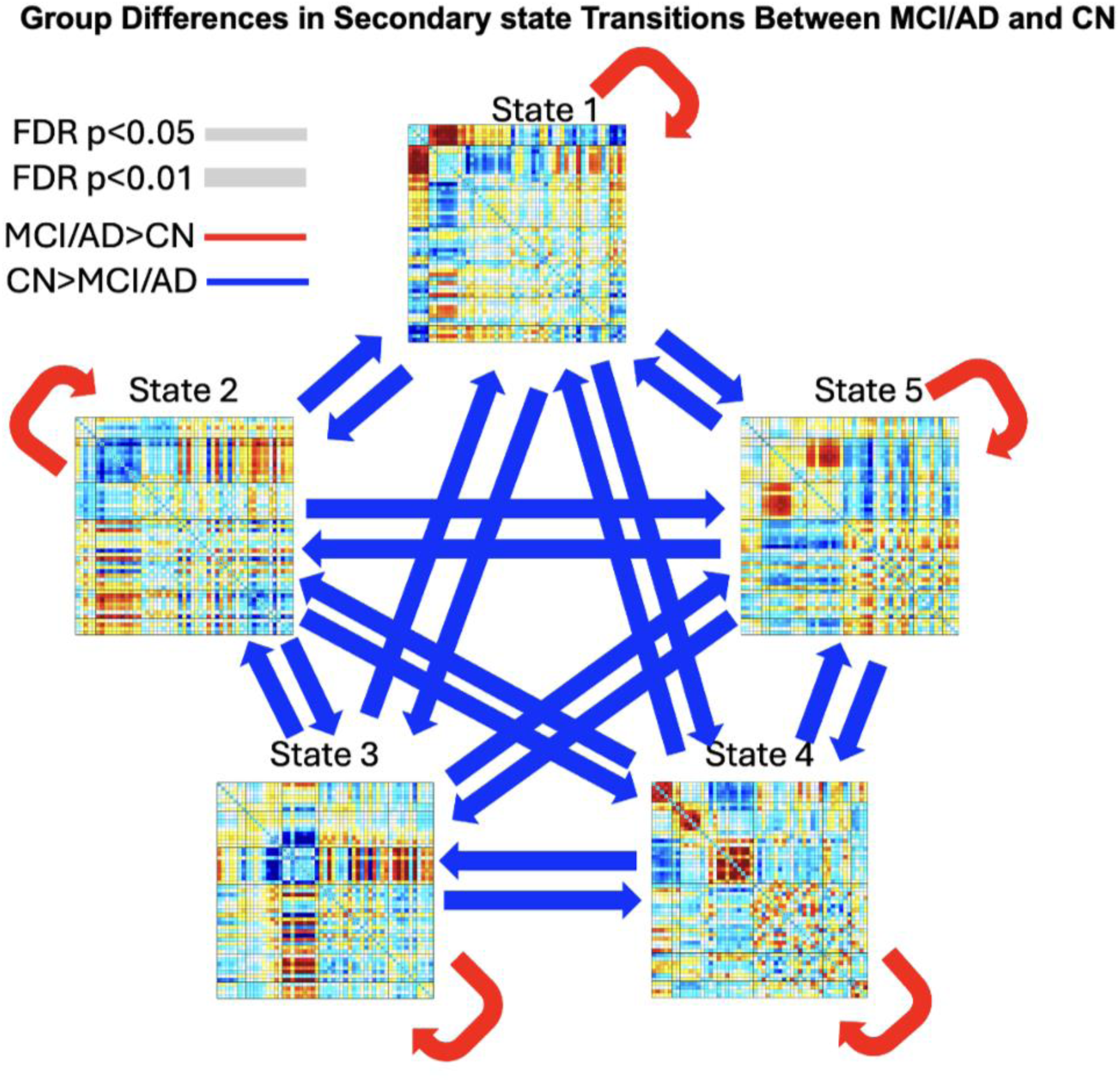
(B) Differences in secondary most probable state transitions among CN, MCI, and AD groups. Arrows indicate significant differences in second-choice transitions. Red: higher in MCI/AD; blue: higher in CN. Arrow thickness reflects group difference magnitude (FDR p < 0.05 or p < 0.01). MCI/AD exhibited more widespread, erratic switching; CN showed stronger self-transitions and more stable engagement with specific states, indicating greater neural stability.

### 3.6. Classification Performance

To assess the discriminative power of dFNC features in classifying MCI/AD patients versus CN individuals, we employed a logistic regression classifier combined with elastic net regularization (ENR) for feature selection, using 5-fold cross-validation (50). A total of 6,960 dynamic features– capturing temporal fluctuations in functional connectivity across brain regions and state transitions–were extracted. These included state-specific connectivity patterns (5 states × 1,378 connections), primary/secondary dwell times, primary/secondary occupancy rates, and primary/secondary state transition probabilities. These features were used to classify MCI/AD subjects against CN. The resulting model achieved a high mean area under the curve (AUC) of approximately 0.89, indicating strong classification performance. To assess generalizability and mitigate potential bias from fixed data splits, we employed a randomized resampling procedure across 10 iterations. In each iteration, a new CN subgroup (n = 689) was randomly sampled from the whole CN cohort to match the number of MCI/AD cases. The model was retrained and re-evaluated in each iteration, and the resulting AUCs were averaged. The mean AUC across iterations was approximately 0.85, suggesting a robust and reproducible classification performance, while also highlighting minor overfitting in the original fixed split.

**Figure 8** presents the top 50 most predictive dFNC features identified by the classification model, mapped onto the five canonical dFNC states. Each circular plot represents one state and displays the most discriminative intra-and inter-network connections. The color bar indicates feature weight direction and magnitude (blue: negative, stronger for CN; red: positive, stronger for MCI/AD), highlighting how specific network interactions contribute to group separation.

**Figure 8.**
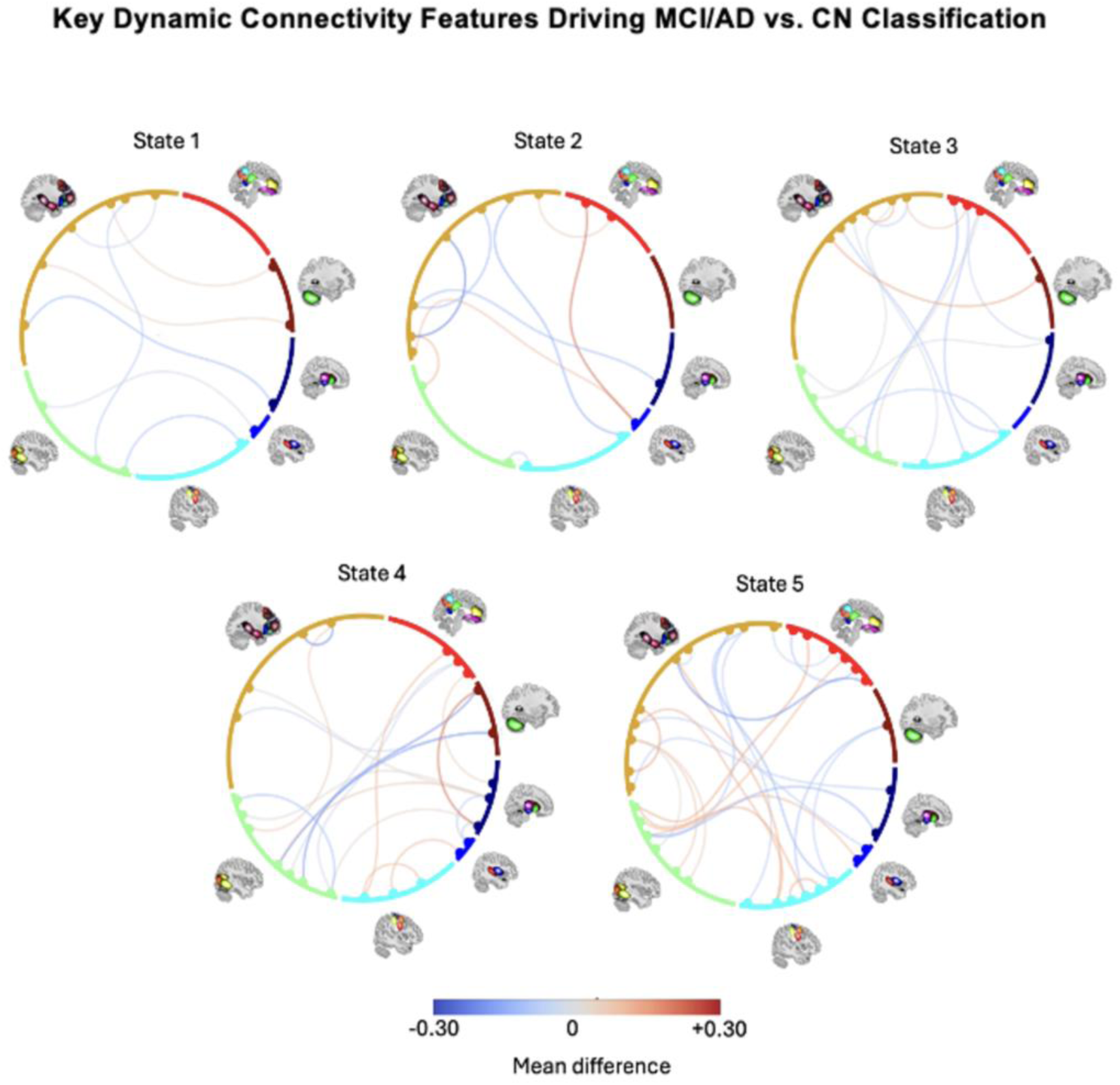
Top 50 most predictive dFNC features for classifying MCI/AD versus CN. Each subplot corresponds to one of the five canonical dFNC states. Connections reflect the most discriminative features selected by Elastic Net Regularization across cross-validation folds. Color intensity represents feature weight (red = MCI/AD, blue = CN). Most high-weight features are in States 1–3 and involve default mode, cognitive control, and subcortical networks—supporting their relevance to neurodegeneration and utility as dFNC biomarkers.

This visualization illustrates both the state-dependent nature of predictive features and the broader functional architecture underlying differences between MCI/AD and CN individuals. The most informative features span across the DM, CC, SM, and SC networks, with dominant patterns concentrated in States 1, 2, and 3–states that also exhibited significant group differences in occupancy and transition probabilities, reinforcing their relevance to AD-related disruptions in brain dynamics.

Our feature selection analysis further revealed a substantial contribution from the CC network in States 1, 2, 3, and 4. The DM network showed consistent involvement in States 3, 4, and 5, particularly through connections with the SM and VIS networks. Additionally, the CB network contributed to classification in all states except State 2, indicating its broad involvement in distinguishing MCI/AD from CN individuals.

The prominence of features within States 1–3, which also differed in occupancy and transition profiles, supports the idea that dynamic states capture meaningful disruptions in network coordination in AD. The alignment between classifier-selected features and altered dynamic states enhances the biological interpretability of our model. Consistent with previous findings (18, 56), which identified the DM and CC networks as key discriminators, our results underscore the value of state-specific dFNC features as potential biomarkers for neurodegeneration.

## 4. Discussion

In this study, we introduced and evaluated a guided dFNC framework using constrained ICA to classify AD and MCI versus CN individuals. By leveraging reference states derived from a large dataset (UKBB) and subsequently constraining ICA in separate cohorts (OASIS-3 and ADNI), our approach provides biologically meaningful priors and robust individual-level dFNC estimates. The key findings from this work, along with their implications, are discussed below.

A significant challenge in dFNC research is the alignment of dFNC states across individuals, especially in heterogeneous populations such as those affected by AD. Traditional approaches often rely on purely data-driven decompositions (e.g., group ICA) and group-level dFNC state estimation, which may not capture the full spectrum of connectivity dynamics relevant to neurodegeneration for any individual (57, 58). By first identifying canonical reference brain states in a large, healthy reference dataset and then guiding ICA in the patient cohorts, we ensured that extracted connectivity patterns would be both reproducible and interpretable at the individual level (26, 59). This method improves upon unconstrained ICA approaches by reducing inter-subject variability, aligning transient states to well-defined FNC “templates,” and enhancing sensitivity to disease-specific changes.

Our results revealed significant differences in dFNC measures between individuals with AD/MCI and those with CN. Across multiple metrics–dwell time, occupancy rate, and transition probability–patients exhibited a general tendency to remain “stuck” in certain connectivity states, reflecting reduced neural flexibility (12, 13, 53, 60, 61). Prolonged engagement in specific states has been associated with a diminished capacity for efficient reconfiguration of brain networks, a phenomenon that may underlie the cognitive and behavioral deficits characteristic of neurodegenerative disorders (21, 57, 62). The observed disruptions involved several large-scale brain networks critical for cognitive function, including default mode, sensorimotor, and cerebellar networks. These findings align with previous reports of altered resting-state network dynamics in AD and MCI, suggesting that compromised switching between connectivity states could serve as an essential biomarker for disease progression (63, 64).

Our analysis went beyond the most dominant (first) connectivity states by also considering the second strongest states, thereby offering a more detailed view of transient connectivity. Interestingly, AD/MCI patients exhibited reduced adaptability not only in their primary states but also in their secondary network states–underscoring the global nature of connectivity disruptions in neurodegeneration (26, 60). This two-tier dwell time analysis revealed that longer dwell times and reduced transition probabilities are widespread, suggesting a breakdown in the flexible re-allocation of neural resources needed for healthy cognitive processing.

By integrating dFNC metrics into a machine learning pipeline that employed logistic regression with elastic net regularization and cross-validated feature learning, we achieved high diagnostic accuracy, with a mean area under the curve (AUC) of approximately 0.85 across repeated sampling. This result underscores the potential of dFNC features as robust biomarkers for early-stage AD(65). As early intervention is essential in AD, approaches that can detect nuanced alterations in resting-state dynamics may contribute to more timely and accurate diagnoses, ultimately improving clinical outcomes (39).

Several methodological innovations underscore the significance of this study. A key advancement is the application of state-wise mean removal, which isolates transient connectivity changes by subtracting stable, long-lasting patterns within each identified state. This enhances sensitivity to subtle dynamic shifts associated with Alzheimer’s disease(57). Another major innovation is the use of template-driven decomposition, in which ICA is constrained using biologically informed priors derived from a large, independent dataset. This approach improves the robustness and interpretability of the decomposition, enhances cross-subject comparability, and addresses limitations inherent in purely data-driven dFNC methods.

Additionally, we introduced a novel set of secondary dFNC features based on the second-most dominant state at each time point, including secondary dwell time, secondary occupancy rate, and secondary state transition probability. These features provide additional insight into subdominant state dynamics that may be critical for detecting early disruptions in functional brain organization(60). Finally, the entire workflow is fully automated, making it scalable and well-suited for extensive neuroimaging studies, where manual processing can be time-intensive and susceptible to observer bias(66).

While this work provides promising insights, several limitations warrant consideration. First, the study datasets originate from multiple sites and scanners, introducing potential confounds due to hardware and acquisition differences. Although we applied standardized preprocessing, further harmonization strategies (e.g., ComBat) could be explored (67–70). Second, resting-state fMRI data inherently suffer from relatively low signal-to-noise ratios and motion artifacts, which can obscure subtle dynamic effects. More advanced denoising strategies or multi-echo acquisitions might further mitigate these confounds (71–73). Additionally, while our classification performance was robust, external validation on entirely independent cohorts and prospective longitudinal data would be valuable to confirm the generalizability and predictive utility of these dynamic features.

A crucial next step will be to link these altered connectivity states with clinical outcomes and longitudinal progression, potentially enabling the development of personalized risk profiles. Future work might also investigate whether combining dFNC features with other imaging modalities (e.g., structural MRI, PET) or fluid biomarkers (e.g., cerebrospinal fluid, blood-based markers) could yield more comprehensive and sensitive diagnostic tools (10, 74).

In summary, our guided dFNC framework, utilizing constrained ICA, successfully identified disease-specific alterations in resting-state dFNC, providing strong evidence for the role of dynamic neural flexibility in AD/MCI. The observed disruptions in dwell time, occupancy rate, and state transition probability support the view that neurodegeneration impairs the brain’s ability to reorganize its connectivity patterns. By introducing a biologically informed, fully automated approach that aligns dFNC states across individuals, this work advances the field of personalized neuroimaging (29). It highlights the potential of dFNC metrics as early markers for AD-related neurodegeneration. Future studies building on this framework may enable more precise tracking of disease trajectories and foster improved interventions for at-risk populations.

Our study builds directly on a series of methodological advances that we previously developed in the area of dynamic functional network connectivity and its clinical applications. This explains the higher proportion of self-citations in the original submission, as many of the analytical approaches employed here are based on our prior work. Nevertheless, we recognize the importance of balancing our references with independent contributions from the broader field, and we have now added multiple external citations to ensure a more comprehensive and representative reference list.

## Code availability

The code used for preprocessing, functional connectivity estimation, and dynamic functional network connectivity (dFNC) calculations is available through the TReNDS software repository (https://trendscenter.org/software/). Statistical Parametric Mapping (SPM12) is openly available at https://www.fil.ion.ucl.ac.uk/spm/. The NeuroMark framework and NeuroMark_fMRI_1.0 template are incorporated into the Group ICA Toolbox (GIFT v4.0.5.14), which is freely available at https://trendscenter.org/software/gift/.

## AI-assisted tools

AI-assisted tools were used only for minor language editing to improve grammar and readability. No AI tools were used for data collection, data analysis, or substantive writing of the manuscript. Software used: Grammarly and ChatGPT for language refinement. Parts of manuscript preparation affected: only the language editing stage. The scientific content, interpretation of results, and conclusions were written and verified solely by the authors. All AI-assisted edits were carefully reviewed and verified by the authors to ensure accuracy, integrity, and compliance with ethical standards.

## Data availability

The UK Biobank (UKBB) imaging dataset used in this manuscript is available through the UK Biobank repository (https://www.ukbiobank.ac.uk/). The Alzheimer’s Disease Neuroimaging Initiative (ADNI) dataset is available upon application at http://adni.loni.usc.edu/. The OASIS-3 dataset is publicly available through the Open Access Series of Imaging Studies (OASIS) project at https://www.oasis-brains.org/. This content is solely the responsibility of the authors and may not reflect the official views of the funders or the original data contributors.

## Author contributions

E.Z., M.S.E.S., A.A., A.I., and V.D.C. contributed to the **conceptualization** of the study, including formulation of the overarching research goals and aims. E.Z. was responsible for **formal analysis, methodology, and visualization**, and prepared the original draft of the manuscript. M.S.E.S. contributed to the **formal analysis and methodology** and was involved in **reviewing and editing** the manuscript. A.A. contributed to **data preprocessing and data collection**. A.I. contributed to the **methodology** and to **reviewing and editing** the manuscript. V.D.C. provided **supervision, conceptual guidance, and funding acquisition**, and contributed to **reviewing and editing** the manuscript. All authors approved the final version of the manuscript.

## Acknowledgements

This work was supported by the US National Institutes of Health (NIH R01AG073949; NIH 1R01AG090597), and the US National Science Foundation (NSF 2316421). Dr. Mohammad S. E. Sendi additionally reports support from the US National Institute of Mental Health (T32MH125786).

## Conflict of Interest

Mohammad Sendi provides consulting services for Niji Corp. All other authors declare no financial interest.

## Notes

### Author Declarations

The Institutional Review Boards of the Alzheimer's Disease Neuroimaging Initiative ADNI, the Open Access Series of Imaging Studies OASIS, the UK Biobank provided ethical approval and obtained informed consent from all participants at their respective sites. The current study involved only secondary analysis of de-identified data, and therefore no additional ethical approval was required.

## References

1. Zhang J, Zhang Y, Wang J, Xia Y, Zhang J, Chen L. Recent advances in Alzheimer’s disease: mechanisms, clinical trials and new drug development strategies. Signal Transduction and Targeted Therapy. 2024;9(1):211.

2. Greicius MD, Srivastava G, Reiss AL, Menon V. Default-mode network activity distinguishes Alzheimer’s disease from healthy aging: evidence from functional MRI. Proc Natl Acad Sci U S A. 2004;101(13):4637–42.

3. Seeley WW, Crawford RK, Zhou J, Miller BL, Greicius MD. Neurodegenerative Diseases Target Large-Scale Human Brain Networks. Neuron. 2009;62(1):42–52.

4. Valera-Bermejo JM, De Marco M, Venneri A. Altered Interplay Among Large-Scale Brain Functional Networks Modulates Multi-Domain Anosognosia in Early Alzheimer’s Disease. Front Aging Neurosci. 2021;13:781465.

5. Albert MS, DeKosky ST, Dickson D, Dubois B, Feldman HH, Fox NC, et al. The diagnosis of mild cognitive impairment due to Alzheimer’s disease: recommendations from the National Institute on Aging-Alzheimer’s Association workgroups on diagnostic guidelines for Alzheimer’s disease. Alzheimers Dement. 2011;7(3):270–9.

6. de Aquino CH. Methodological Issues in Randomized Clinical Trials for Prodromal Alzheimer’s and Parkinson’s Disease. Front Neurol. 2021;12:694329.

7. Chouliaras L, O’Brien JT. The use of neuroimaging techniques in the early and differential diagnosis of dementia. Molecular Psychiatry. 2023;28(10):4084–97.

8. Schindler SE, Galasko D, Pereira AC, Rabinovici GD, Salloway S, Suárez-Calvet M, et al. Acceptable performance of blood biomarker tests of amyloid pathology — recommendations from the Global CEO Initiative on Alzheimer’s Disease. Nature Reviews Neurology. 2024;20(7):426–39.

9. Grøntvedt GR, Lauridsen C, Berge G, White LR, Salvesen Ø, Bråthen G, et al. The Amyloid, Tau, and Neurodegeneration (A/T/N) Classification Applied to a Clinical Research Cohort with Long-Term Follow-Up. J Alzheimers Dis. 2020;74(3):829–37.

10. Jack CR, Jr., Bennett DA, Blennow K, Carrillo MC, Dunn B, Haeberlein SB, et al. NIA-AA Research Framework: Toward a biological definition of Alzheimer’s disease. Alzheimers Dement. 2018;14(4):535–62.

11. Pini L, Lista S, Griffa A, Allali G, Imbimbo BP. Can brain network connectivity facilitate the clinical development of disease-modifying anti-Alzheimer drugs? Brain Communications. 2024;7(1).

12. Córdova-Palomera A, Kaufmann T, Persson K, Alnæs D, Doan NT, Moberget T, et al. Disrupted global metastability and static and dynamic brain connectivity across individuals in the Alzheimer’s disease continuum. Scientific Reports. 2017;7(1):40268.

13. Schumacher J, Peraza LR, Firbank M, Thomas AJ, Kaiser M, Gallagher P, et al. Dynamic functional connectivity changes in dementia with Lewy bodies and Alzheimer’s disease. NeuroImage: Clinical. 2019;22:101812.

14. Biswal B, Yetkin FZ, Haughton VM, Hyde JS. Functional connectivity in the motor cortex of resting human brain using echo-planar MRI. Magn Reson Med. 1995;34(4):537–41.

15. Whitten LA. RTI Press Occasional Papers. Functional Magnetic Resonance Imaging (fMRI): An Invaluable Tool in Translational Neuroscience. Research Triangle Park (NC): RTI Press © 2012 Research Triangle Institute. All rights reserved.; 2012.

16. Tang X, Wang L, Feng Q, Hu H, Zhu Y, Liao Z, et al. Resting-state functional magnetic resonance imaging study on cerebrovascular reactivity changes in the precuneus of Alzheimer’s disease and mild cognitive impairment patients. Scientific Reports. 2025;15(1):363.

17. Damoiseaux JS. Resting-state fMRI as a biomarker for Alzheimer’s disease? Alzheimers Res Ther. 2012;4(2):8.

18. Gu Y, Lin Y, Huang L, Ma J, Zhang J, Xiao Y, et al. Abnormal dynamic functional connectivity in Alzheimer’s disease. CNS Neurosci Ther. 2020;26(9):962–71.

19. Canal-Garcia A, Veréb D, Mijalkov M, Westman E, Volpe G, Pereira JB. Dynamic multilayer functional connectivity detects preclinical and clinical Alzheimer’s disease. Cereb Cortex. 2024;34(2).

20. Chang C, Glover GH. Time-frequency dynamics of resting-state brain connectivity measured with fMRI. Neuroimage. 2010;50(1):81–98.

21. Calhoun Vince D, Miller R, Pearlson G, Adalı T. The Chronnectome: Time-Varying Connectivity Networks as the Next Frontier in fMRI Data Discovery. Neuron. 2014;84(2):262–74.

22. Hutchison RM, Womelsdorf T, Allen EA, Bandettini PA, Calhoun VD, Corbetta M, et al. Dynamic functional connectivity: Promise, issues, and interpretations. NeuroImage. 2013;80:360–78.

23. Shine James M, Bissett Patrick G, Bell Peter T, Koyejo O, Balsters Joshua H, Gorgolewski Krzysztof J, et al. The Dynamics of Functional Brain Networks: Integrated Network States during Cognitive Task Performance. Neuron. 2016;92(2):544–54.

24. Hutchison RM, Morton JB. Tracking the Brain’s Functional Coupling Dynamics over Development. J Neurosci. 2015;35(17):6849–59.

25. de Vos F, Koini M, Schouten TM, Seiler S, van der Grond J, Lechner A, et al. A comprehensive analysis of resting state fMRI measures to classify individual patients with Alzheimer’s disease. Neuroimage. 2018;167:62–72.

26. Fu Z, Caprihan A, Chen J, Du Y, Adair JC, Sui J, et al. Altered static and dynamic functional network connectivity in Alzheimer’s disease and subcortical ischemic vascular disease: shared and specific brain connectivity abnormalities. Hum Brain Mapp. 2019;40(11):3203–21.

27. Jing R, Chen P, Wei Y, Si J, Zhou Y, Wang D, et al. Altered large-scale dynamic connectivity patterns in Alzheimer’s disease and mild cognitive impairment patients: A machine learning study. Hum Brain Mapp. 2023;44(9):3467–80.

28. Du Y, Fan Y. Group information guided ICA for fMRI data analysis. Neuroimage. 2013;69:157–97.

29. Finn ES, Shen X, Scheinost D, Rosenberg MD, Huang J, Chun MM, et al. Functional connectome fingerprinting: identifying individuals using patterns of brain connectivity. Nature Neuroscience. 2015;18(11):1664–71.

30. Preti MG, Bolton TA, Van De Ville D. The dynamic functional connectome: State-of-the-art and perspectives. Neuroimage. 2017;160:41–54.

31. Abrol A, Fu Z, Du Y, Calhoun V. Multimodal Data Fusion of Deep Learning and Dynamic Functional Connectivity Features to Predict Alzheimer’s Disease Progression *2019. 4409–13 p.

32. Lin QH, Liu J, Zheng YR, Liang H, Calhoun VD. Semiblind spatial ICA of fMRI using spatial constraints. Hum Brain Mapp. 2010;31(7):1076–88.

33. Du Y, Fu Z, Sui J, Gao S, Xing Y, Lin D, et al. NeuroMark: An automated and adaptive ICA based pipeline to identify reproducible fMRI markers of brain disorders. NeuroImage: Clinical. 2020;28:102375.

34. Iraji A, Fu Z, Faghiri A, Duda M, Chen J, Rachakonda S, et al. Identifying canonical and replicable multi-scale intrinsic connectivity networks in 100k+ resting-state fMRI datasets. Hum Brain Mapp. 2023;44(17):5729–48.

35. Long Z, Wang Z, Zhang J, Zhao X, Yao L. Temporally constrained ICA with threshold and its application to fMRI data. BMC Medical Imaging. 2019;19(1):6.

36. Khalilullah KMI, Agcaoglu O, Sui J, Duda M, Adali T, Calhoun VD. Parallel Multilink Group Joint ICA: Fusion of 3D Structural and 4D Functional Data Across Multiple Resting fMRI Networks. bioRxiv. 2024.

37. Miller KL, Alfaro-Almagro F, Bangerter NK, Thomas DL, Yacoub E, Xu J, et al. Multimodal population brain imaging in the UK Biobank prospective epidemiological study. Nature Neuroscience. 2016;19(11):1523–36.

38. LaMontagne PJ, Benzinger TL, Morris JC, Keefe S, Hornbeck R, Xiong C, et al. OASIS-3: Longitudinal Neuroimaging, Clinical, and Cognitive Dataset for Normal Aging and Alzheimer Disease. medRxiv. 2019:2019.12.13.19014902.

39. Jack CR, Jr., Bernstein MA, Fox NC, Thompson P, Alexander G, Harvey D, et al. The Alzheimer’s Disease Neuroimaging Initiative (ADNI): MRI methods. J Magn Reson Imaging. 2008;27(4):685–91.

40. Hyman BT, Phelps CH, Beach TG, Bigio EH, Cairns NJ, Carrillo MC, et al. National Institute on Aging-Alzheimer’s Association guidelines for the neuropathologic assessment of Alzheimer’s disease. Alzheimers Dement. 2012;8(1):1–13.

41. Morris JC. The Clinical Dementia Rating (CDR). Neurology. 1993;43(11):2412--a.

42. Calhoun VD, Adali T, Pearlson GD, Pekar JJ. A method for making group inferences from functional MRI data using independent component analysis. Hum Brain Mapp. 2001;14(3):140–51.

43. Dini H, Bruni LE, Ramsøy TZ, Calhoun VD, Sendi MSE. The overlap across psychotic disorders: A functional network connectivity analysis. International Journal of Psychophysiology. 2024;201:112354.

44. Sendi MSE, Fu Z, Harnett NG, van Rooij SJH, Vergara V, Pizzagalli DA, et al. Brain dynamics reflecting an intra-network brain state are associated with increased post-traumatic stress symptoms in the early aftermath of trauma. Nature Mental Health. 2025;3(2):185–98.

45. Fu Z, Abbott CC, Miller J, Deng Z-D, McClintock SM, Sendi MSE, et al. Cerebro-cerebellar functional neuroplasticity mediates the effect of electric field on electroconvulsive therapy outcomes. Translational Psychiatry. 2023;13(1):43.

46. Allen EA, Damaraju E, Plis SM, Erhardt EB, Eichele T, Calhoun VD. Tracking Whole-Brain Connectivity Dynamics in the Resting State. Cerebral Cortex. 2012;24(3):663–76.

47. Soleimani N, Iraji A, Pearlson G, Preda A, Calhoun VD. Unraveling the Neural Landscape of Mental Disorders using Double Functional Independent Primitives (dFIPs). Biol Psychiatry Cogn Neurosci Neuroimaging. 2025.

48. Damaraju E, Allen EA, Belger A, Ford JM, McEwen S, Mathalon DH, et al. Dynamic functional connectivity analysis reveals transient states of dysconnectivity in schizophrenia. NeuroImage: Clinical. 2014;5:298–308.

49. Vidaurre D, Smith SM, Woolrich MW. Brain network dynamics are hierarchically organized in time. Proc Natl Acad Sci U S A. 2017;114(48):12827–32.

50. Zou H, Hastie T. Regularization and Variable Selection Via the Elastic Net. Journal of the Royal Statistical Society Series B: Statistical Methodology. 2005;67(2):301–20.

51. Jo T, Nho K, Saykin AJ. Deep Learning in Alzheimer’s Disease: Diagnostic Classification and Prognostic Prediction Using Neuroimaging Data. Front Aging Neurosci. 2019;11:220.

52. Prvulovic D, Bokde ALW, Faltraco F, Hampel H. Functional magnetic resonance imaging as a dynamic candidate biomarker for Alzheimer’s disease. Progress in Neurobiology. 2011;95(4):557–69.

53. Sendi MSE, Zendehrouh E, Fu Z, Liu J, Du Y, Mormino E, et al. Disrupted Dynamic Functional Network Connectivity Among Cognitive Control Networks in the Progression of Alzheimer’s Disease. Brain Connect. 2023;13(6):334–43.

54. Meng X, Iraji A, Fu Z, Kochunov P, Belger A, Ford JM, et al. Multi-model order spatially constrained ICA reveals highly replicable group differences and consistent predictive results from resting data: A large N fMRI schizophrenia study. Neuroimage Clin. 2023;38:103434.

55. Sendi MSE, Zendehrouh E, Miller RL, Fu Z, Du Y, Liu J, et al. Alzheimer’s Disease Projection From Normal to Mild Dementia Reflected in Functional Network Connectivity: A Longitudinal Study. Front Neural Circuits. 2020;14:593263.

56. Khatri U, Kwon G-R. Classification of Alzheimer’s Disease and Mild-Cognitive Impairment Base on High-Order Dynamic Functional Connectivity at Different Frequency Band. Mathematics. 2022;10(5):805.

57. Lurie DJ, Kessler D, Bassett DS, Betzel RF, Breakspear M, Kheilholz S, et al. Questions and controversies in the study of time-varying functional connectivity in resting fMRI. Network Neuroscience. 2020;4(1):30–69.

58. Leonardi N, Van De Ville D. On spurious and real fluctuations of dynamic functional connectivity during rest. NeuroImage. 2015;104:430–6.

59. Vidaurre D, Abeysuriya R, Becker R, Quinn AJ, Alfaro-Almagro F, Smith SM, et al. Discovering dynamic brain networks from big data in rest and task. NeuroImage. 2018;180:646–56.

60. Viviano RP, Raz N, Yuan P, Damoiseaux JS. Associations between dynamic functional connectivity and age, metabolic risk, and cognitive performance. Neurobiology of Aging. 2017;59:135–43.

61. Zalesky A, Fornito A, Cocchi L, Gollo LL, Breakspear M. Time-resolved resting-state brain networks. Proceedings of the National Academy of Sciences. 2014;111(28):10341–6.

62. Bassett DS, Sporns O. Network neuroscience. Nature Neuroscience. 2017;20(3):353–64.

63. Zhou J, Greicius MD, Gennatas ED, Growdon ME, Jang JY, Rabinovici GD, et al. Divergent network connectivity changes in behavioural variant frontotemporal dementia and Alzheimer’s disease. Brain. 2010;133(Pt 5):1352–67.

64. Jones DT, Vemuri P, Murphy MC, Gunter JL, Senjem ML, Machulda MM, et al. Non-stationarity in the “resting brain’s” modular architecture. PLoS One. 2012;7(6):e39731.

65. Suk H-I, Lee S-W, Shen D. Hierarchical feature representation and multimodal fusion with deep learning for AD/MCI diagnosis. NeuroImage. 2014;101:569–82.

66. Casey BJ, Cannonier T, Conley MI, Cohen AO, Barch DM, Heitzeg MM, et al. The Adolescent Brain Cognitive Development (ABCD) study: Imaging acquisition across 21 sites. Developmental Cognitive Neuroscience. 2018;32:43–54.

67. Albers AM, Meindertsma T, Toni I, de Lange FP. Decoupling of BOLD amplitude and pattern classification of orientation-selective activity in human visual cortex. NeuroImage. 2018;180:31–40.

68. Fortin J-P, Parker D, Tunç B, Watanabe T, Elliott MA, Ruparel K, et al. Harmonization of multi-site diffusion tensor imaging data. NeuroImage. 2017;161:149–70.

69. Gardner M, Shinohara RT, Bethlehem RAI, Romero-Garcia R, Warrier V, Dorfschmidt L, et al. ComBatLS: A Location-and Scale-Preserving Method for Multi-Site Image Harmonization. Hum Brain Mapp. 2025;46(8):e70197.

70. Yamashita A, Yahata N, Itahashi T, Lisi G, Yamada T, Ichikawa N, et al. Harmonization of resting-state functional MRI data across multiple imaging sites via the separation of site differences into sampling bias and measurement bias. PLoS Biol. 2019;17(4):e3000042.

71. Kundu P, Inati SJ, Evans JW, Luh WM, Bandettini PA. Differentiating BOLD and non-BOLD signals in fMRI time series using multi-echo EPI. Neuroimage. 2012;60(3):1759–70.

72. Cohen AD, Yang B, Fernandez B, Banerjee S, Wang Y. Improved resting state functional connectivity sensitivity and reproducibility using a multiband multi-echo acquisition. Neuroimage. 2021;225:117461.

73. Dipasquale O, Sethi A, Laganà MM, Baglio F, Baselli G, Kundu P, et al. Comparing resting state fMRI de-noising approaches using multi-and single-echo acquisitions. PLoS One. 2017;12(3):e0173289.

74. Jack CR, Knopman DS, Jagust WJ, Petersen RC, Weiner MW, Aisen PS, et al. Tracking pathophysiological processes in Alzheimer’s disease: an updated hypothetical model of dynamic biomarkers. The Lancet Neurology. 2013;12(2):207–16.

